# Social Determinants of Genetics Referral and Completion Rates Among Child Neurology Patients

**DOI:** 10.1101/2023.09.12.23295450

**Authors:** Jordan J. Cole, Angela D. Sellitto, Laura Rosa Baratta, Julia B. Huecker, Joyce E. Balls-Berry, Christina A. Gurnett

## Abstract

**Objective:** To investigate clinical, social, and systems-level determinants predictive of genetics clinic referral and completion of genetics clinic visits among child neurology patients.

**Methods:** Electronic health record data were extracted from patients 0-18 years old who were evaluated in child neurology clinics at a single tertiary care institution between July 2018 to January 2020. Variables aligned with the Health Equity Implementation Framework. Referral and referral completion rates to genetics and cardiology clinics were compared among Black vs White patients using bivariate analysis. Demographic variables associated with genetics clinic referral and visit completion were identified using logistic regressions.

**Results:** In a cohort of 11,371 child neurology patients, 304 genetics clinic referrals and 82 cardiology clinic referrals were placed. In multivariate analysis of patients with Black or White ethnoracial identity (n=10,601), genetics clinic referral rates did not differ by race, but were significantly associated with younger age, rural address, neurodevelopmental disorder diagnosis, number of neurology clinic visits, and provider type. The only predictors of genetics clinic visit completion number of neurology clinic visits and race/ethnicity, with White patients being twice as likely as Black patients to complete the visit. Cardiology clinic referrals and visit completion did not differ by race/ethnicity.

**Interpretation:** Although race/ethnicity was not associated with differences in genetics clinic referral rates, White patients were twice as likely as Black patients to complete a genetics clinic visit after referral. Further work is needed to determine whether this is due to systemic/structural racism, differences in attitudes toward genetic testing, or other factors.

## Introduction

Genetic testing is an increasingly common component to the practice of pediatric neurology in the United States (US). The American College of Medical Genetics (ACMG) recommends exome or genome sequencing (ES/GS) as first or second line testing in the diagnostic evaluation of global developmental delay/intellectual disability (GDD/ID),^1^ and the American Epilepsy Society (AES) and National Society of Genetic Counselors recommend ES/GS or a multi-gene panel (MGP) as first-tier testing for unexplained epilepsy.^2^ In these populations, the diagnostic yield of ES/GS is up to 40%,^3,4^ and benefits of testing include specific changes in medical management, surveillance, prognosis, family-planning, and/or access to research or support groups.^3,5-7^ ES/GS has been found to have similar yield and benefits among individuals with other pediatric-onset neurologic conditions including cerebral palsy, neuromuscular disorders, and microcephaly.^8,9,10^ Though ES/GS have the highest diagnostic yield compared to other genetic tests,^11,12^ they are often not easily accessible due to barriers including insurance coverage, out of pocket cost, and access to specialists/genetic counselors.^13-16^ In many cases, referral to genetics specialists is required for patients to obtain ES/GS.^16^ Other types of genetic testing such as chromosomal microarray (CMA) or single-gene testing may be more easily accessible but can still present challenges such as interpretation of variants of unknown significance (VUS) and family counseling.^17^

While these barriers can affect all patients, patients from historically marginalized groups may be disproportionately affected,^18,19^ and often face additional barriers including the effects of structural racism, discrimination, implicit bias, and medical mistrust due to historical injustices in the U.S. health system.^20-22^ Prior studies have found that children identified as Black/African American (Black/AA) or Hispanic are significantly less likely to receive genetic testing compared to non-Hispanic White children,^23-25^ but the relationship between race/ethnicity and other social, clinical, and systems level determinants has not been previously explored. We used an implementation science framework to evaluate the barriers and facilitators to equitable genetic testing among children evaluated in our institution’s child neurology clinics. This work is part of a larger body of ongoing research exploring the diagnostic pathways for genetic testing at tertiary care children’s hospitals. This paper assesses the predictors of referral to genetics clinic referral and genetics visit completion rates of child neurology patients at Washington University in St. Louis (WUSTL). We aimed to determine whether clinical, social, and/or systems-level determinants are predictive of access to genetic services so that we could identify targets for potential intervention to reduce disparities if they are present.

## Methods

Retrospective electronic health record (EHR) data were extracted for all patients evaluated in child neurology clinic at a single tertiary care institution (Washington University in St. Louis or WUSTL) between July 1, 2018 to January 1, 2020. The Health Equity Implementation Framework^26^ was chosen as a model to guide study design in terms of the independent variables to be assessed. This is a determinant framework, meaning the goal is to establish the factors that predict successful implementation of the innovation to be studied, in this case genetic service usage. It is essential to consider contextual factors at both the societal and organizational/local levels, as well as individual-level factors pertaining to patients and providers (**Figure 1**).

**Figure 1.**
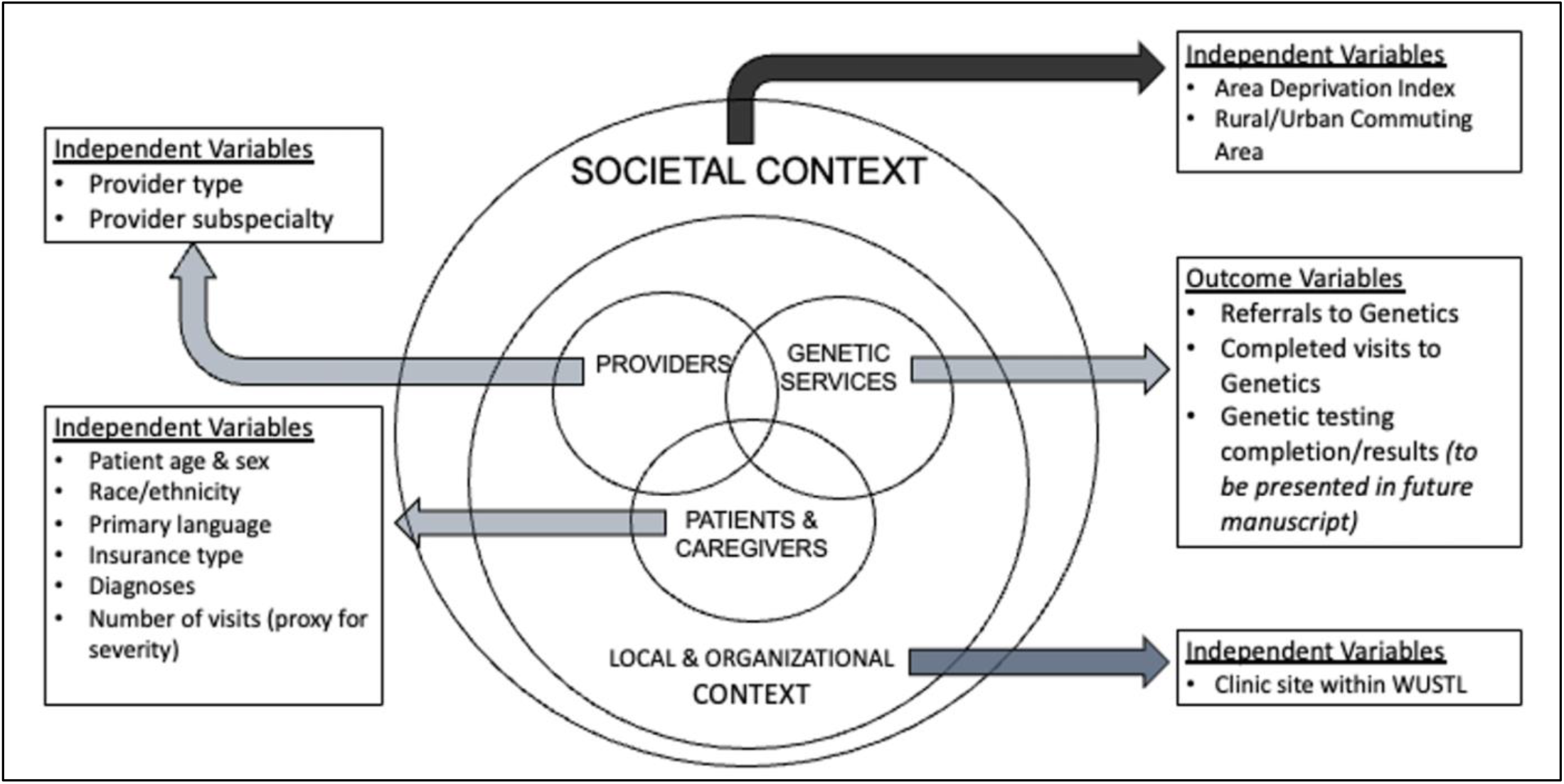
Alignment of Study Variables with the Health Equity Implementation Framework: The independent variables included in this study of genetic service access were aligned across multiple levels of this simplified version of the Health Equity Implementation Framework.^26^ Deeper evaluation of provider and patient-level factors including motivation, genetic knowledge, and bias/discrimination is beyond the scope of this EHR study but planned for future mixed methods work.

### Current clinical workflow

The process for obtaining genetic testing for all children evaluated in Child Neurology Clinics during the study dates is outlined in **Figure 2**. This was determined through the authors’ own clinical experience as well as discussions with clinic leadership.

**Figure 2.**
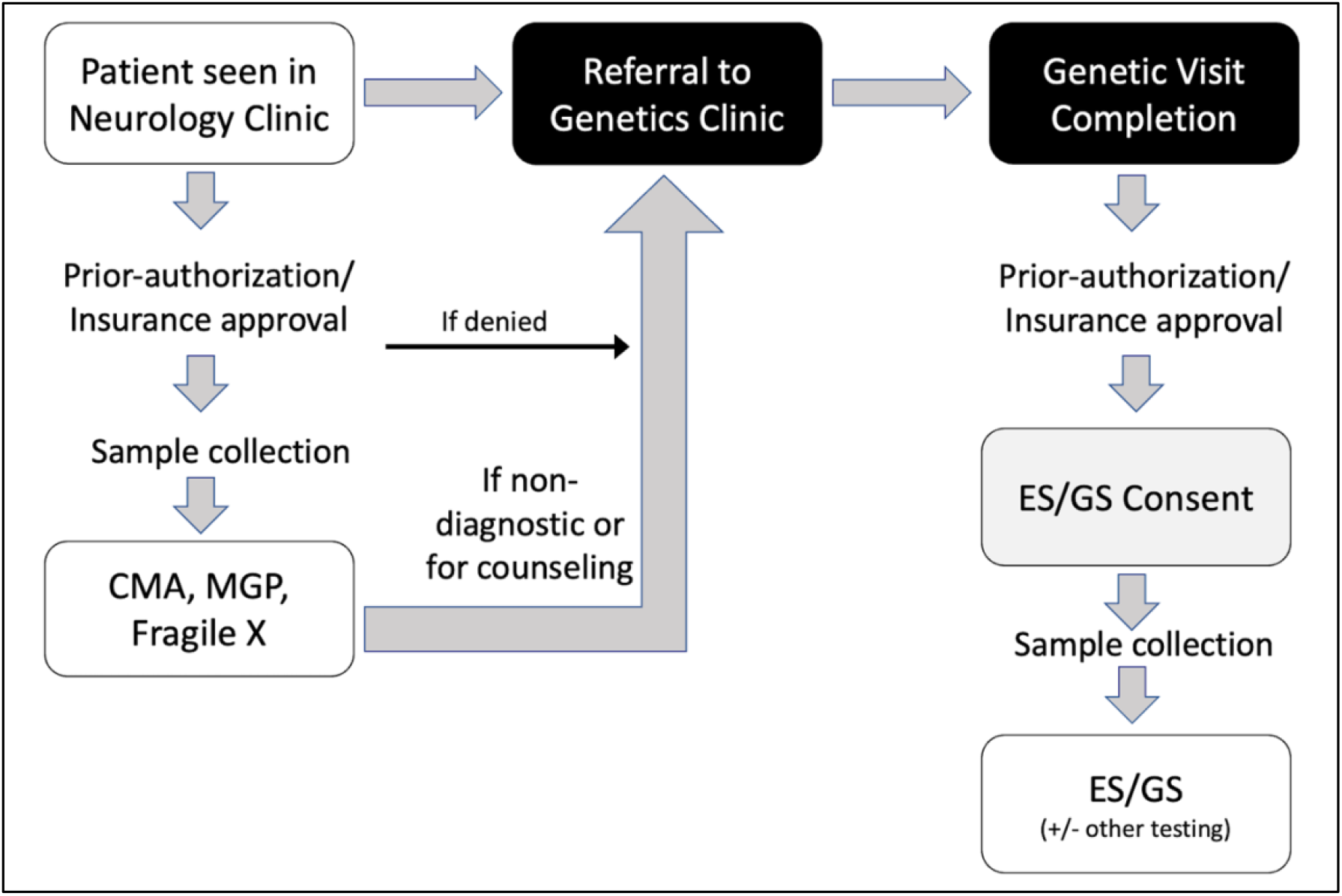
Genetic Testing Workflow at Child Neurology Clinics: Patients seen in child neurology clinics at WUSTL may have some genetic tests requested/ordered directly by their neurologist, such as chromosomal microarray (CMA), multigene panels (MGP), and fragile X testing. However, exome/genome sequencing (ES/GS), requires a referral to Genetics Clinic.

### Demographic variables

Race/ethnicity were combined into a single category given that both are social constructs without a biological basis.^27^ The purpose of racial/ethnic categorization in this study was to assess inequities in access to care on the basis of social factors. Race/ethnicity as documented in the EHR was either caregiver/self-reported or entered by clinical staff; rates of each are not known as this is not recorded. Biological sex was assessed due to the higher rates of neurodevelopmental disorders (NDD) among males.^28^ Age (in months) was recorded at the first neurology clinic visit within the study dates. Primary language was categorized as English or non-English. Socioeconomic advantage/disadvantage was assessed through the Area Deprivation Index (ADI), a validated measure that incorporates data from the U.S. Census and American Community Survey including poverty rates, education, housing, and employment.^29^ ADI was obtained for each individual through geocoding addresses to the Census block group level, through 9-digit-zipcode mapping, or (when neither available) through manual address entry on the ADI website, which produces the ADI at the smallest geographic level available.^30^ Rural/urban status was assessed through the Rural Urban Commuting Area (RUCA), a measure of population density, urbanization, and daily commuting created by the U.S. Department of Agriculture.^31^ RUCA was obtained through geocoding to the Census tract level or (when unavailable) 5-digit-zipcode mapping. Results were dichotomized to rural or urban per the Rural Health Research Center Classification C.^32^ Insurance status was categorized as private, public, or other/uninsured, which included Tricare.

### Clinical variables

Provider level of training and sub-specialization status were combined into a single variable. Patients were categorized based on the highest level of training by any of the providers they saw during the study dates. Site seen within WUSTL accounted for all locations at which the patient was seen during the study timeframe (St. Louis Children’s Hospital [SLCH] Main Campus, Children’s Specialty Care Center [CSCC], other WUSTL site, or a combination of the three). International Classification of Diseases, Tenth Revision, Clinical Modification (ICD-10-CM) codes and Systematized Nomenclature of Medicine Clinical Terms (SNOMED-CT) codes were obtained for all patient visits. SNOMED-CT codes were mapped to ICD-10-CM codes using the March 2019 National Library of Medicine standardized mapping tool.^33^ The Clinical Classifications Software Refined (CCSR) database was then used to identify patients with NDD, seizures/epilepsy, and headache/migraine.^34^ Number of neurology visits within the study time frame was measured as a proxy for severity of illness.

### Outcomes variables

EHR orders for referrals to genetics placed by neurology were extracted and completion status of referral orders was manually confirmed through chart review. Genetics referrals were considered completed if a patient was seen in genetics clinic within one year of the referral order. As a control comparison, referral orders to cardiology and completion status of cardiology visits after referral were also measured.

### Statistical analysis

Bivariate comparisons of demographic and clinical features between Black and White patients were completed using Chi-square tests. P-value <0.05 were considered statistically significant. Univariate logistic regression models were used to predict which patients were referred to genetics clinic using Proc Logistic of SAS software version 9.4.^35^ For unordered categorical predictors, odd ratio (ORs) were used to compare to the referent category. For ordered predictors, ORs were expressed for a one-unit increase in the predictor, except where noted. Due to skewed distributions and/or poor model fit, inherently continuous variables were categorized for analyses. We *a priori* defined quintiles categories of ADI percentile based on ADI National quintiles. P-value <0.05 were considered statistically significant. Diagnostics of collinearity was performed by linear regression; lack of collinearity was verified when the variance inflation factor was 2 or less.^36^ Multivariable logistic regression models were built using the forward selection option with an entry p-value of 0.1 and to stay in the model a p-value of <0.1.

The study was approved by the WUSTL Institutional Review Board (IRB) as exempt per 2018 Common Rule Exempt Categories (45 CFR 46.104). Informed consent was not required for this retrospective review study.

## Results

### Clinic Population Demographics & Clinical Features

A total of 11,371 unique patients were seen at a total of 19,177 child neurology clinic visits (mean of 1.69 visits per patient, SD 0.89, range 1-13) between July 1, 2018 to January 1, 2020. The average patient age at their first visit within the study timeframe was 111 months (9.25 years; SD 64.2 months). The racial/ethnic breakdown of all child neurology clinic patients was 78.1% non-Hispanic White, 14.6% non-Hispanic Black/AA, 3.0% Hispanic, 2.4% non-Hispanic Other, 0.7% non-Hispanic Multiracial, and 1.2% Unknown/Declined to report. The 2020 U.S. Census racial/ethnic demographics of the St. Louis Missouri-Illinois (MO-IL) metropolitan area^37^ and the state of Missouri^38^, which make up a large proportion of the catchment area for the clinic, were, respectively, 70.3% and 75.8% White, 17.8% and 11.3% Black/AA, 3.8% and 4.9% Hispanic, 3.5% and 3.0% Other, and 4.5% and 5% Multiracial. Additional demographic, clinical, and visit-related features of the total clinic population are provided in **Table 1** with a comparison among patients identified as White (non-Hispanic, non-multiracial) and those identified as Black (non-Hispanic, including multiracial where Black/AA was one of the selected races).

**Table 1:**
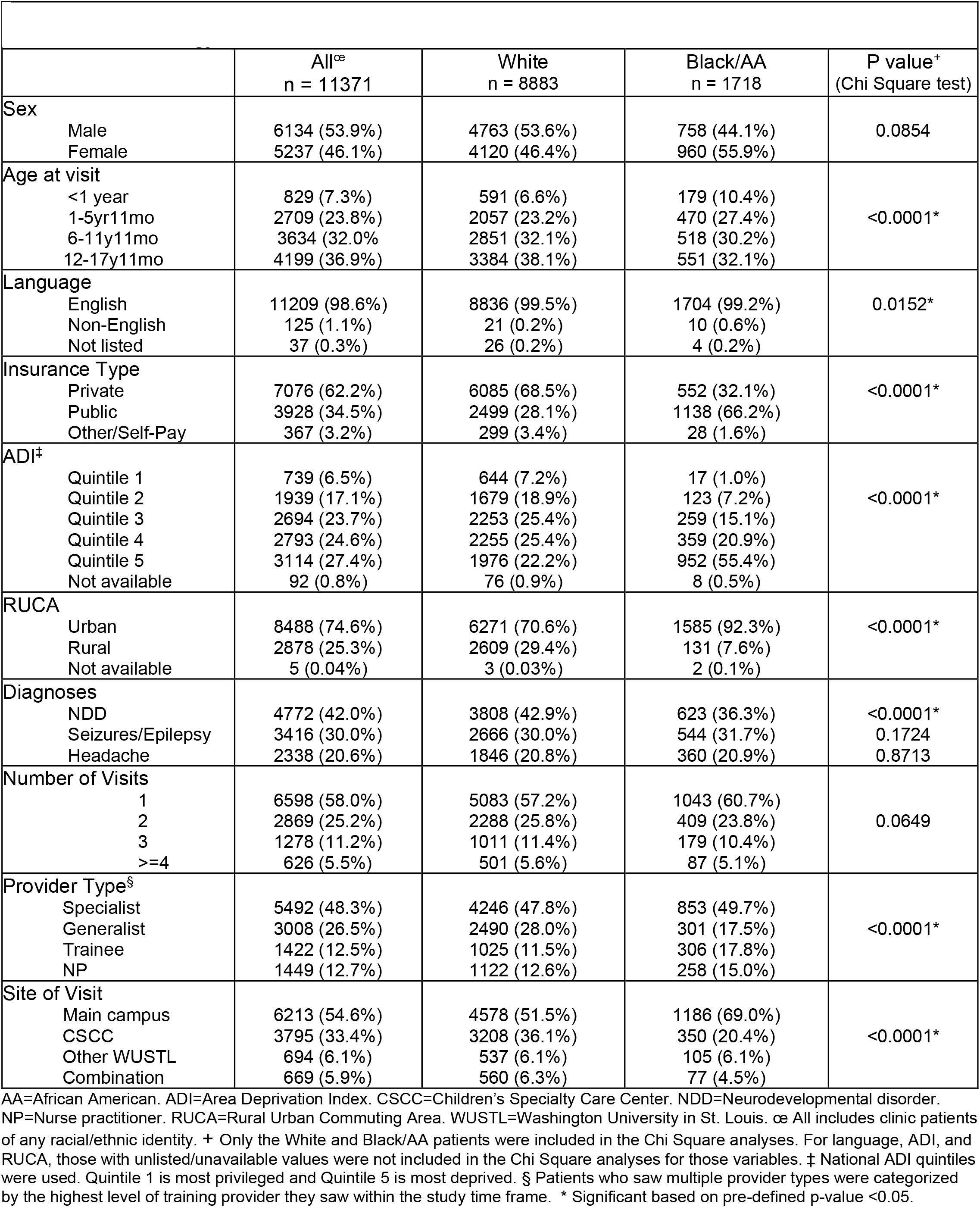
Comparison of Demographic and Clinical Features Between Black/African American and White Child Neurology Clinic Patients.

### Genetic & Cardiology Referrals and Completion Status

Of 11,371 patients seen in child neurology clinics, 304 genetics clinic referrals and 82 cardiology clinic referrals were placed. There was no statistical difference in the percentage of patients referred to genetics clinic among White (2.7%) and Black patients (2.4%) (p=0.442) (**Table 2**). However, significantly fewer Black patients completed the genetics clinic visit compared to White patients (63.4% vs. 78.0%) (p=0.046). There were no differences in the cardiology clinic referral or visit completion rates among Black and White patients.

**Table 2:** Genetic & Cardiology Referrals and Completion Status Between Black and White Patients.

|  | White<br>N (%) | Black/AA<br>N (%) | P value<sup>‡</sup> | OR (95% CI)<sup>§</sup> |
| --- | --- | --- | --- | --- |
| Genetics Referral |  |  |  |  |
| Yes | 241 (2.7%) | 41 (2.4%) |  |  |
| No | 8642 (97.3%) | 1677 (97.6%) | 0.4417 | 1.14 (0.82-1.59) |
| Genetics Visit Completed |  |  |  |  |
| Yes | 188 (78.0%) | 26 (63.4%) |  |  |
| No | 53 (22.0%) | 15 (36.6%) | 0.0464* | 2.05 (1.01-4.14) |
| Cardiology Referral |  |  |  |  |
| Yes | 65 (0.7%) | 10 (0.6%) |  |  |
| No | 8818 (99.3%) | 1708 (99.4%) | 0.4993 | 1.26 (0.65-2.45) |
| Cardiology Visit Completed |  |  |  |  |
| Yes | 53 (81.5%) | 10 (100%) |  |  |
| No | 12 (18.5%) | 0 (0%) | 0.3490 | N/A |

AA=African American. N/A=Not applicable. ‡ P values are from univariate logistic regression except for Cardiology Visit Completed, which used Chi Square because all 10 Black/AA patients completed the visits and thus odds ratio could not be computed for race. § OR=Odds ratio and CI=Confidence interval, reported for White race; Black race was the reference category. \* Significant based on pre-defined p-value <0.05.

### Predictors of Genetics Referral Status in Univariate and Multivariate Analysis

To assess predictors of referral to genetics clinic, univariate analysis was completed for 10,601 patients identified as non-Hispanic Black (including multiracial) or non-Hispanic White who were evaluated in child neurology clinic. Thirteen independent variables representing social, clinical, and systems-level determinants of health were assessed **(Table 3)**. On univariate analysis, there were 9 significant positive predictors of genetics referral including male sex, younger age, public insurance, higher ADI quintile (more socioeconomic deprivation), rural address, NDD diagnosis, higher number of neurology visits, specialist or trainee provider type, and clinic site. There were 3 significant negative predictors of genetics clinic referral which were headache/migraine diagnosis, nurse practitioner (NP)-only provider type, and being seen at the CSCC clinic site. Variables with p-values <0.1 were subsequently assessed as candidate predictors in the multivariate regression model. Ultimately, 5 predictors of genetic referral remained statistically significant in the final multivariate model, including younger age, rural address, NDD diagnosis, number of visits, and provider type.

**Table 3:** Association of Demographic and Clinical Variables with Genetics Clinic Referral Status.

| Predictor<sup>+</sup> | Univariate OR<br>(95% CI) | P value | Multivariate OR<sup>£</sup><br>(95% CI) | P value |
| --- | --- | --- | --- | --- |
| Male | 1.32 (1.04-1.69) | 0.0235* |  |  |
| Age Category<sup>§</sup> | 0.539 (0.48-0.61) | <0.0001* | 0.61 (0.54-0.69) | <0.001* |
| White race | 1.14 (0.82-1.59) | 0.4417 |  |  |
| Non-English language | 2.53 (0.6-10.6) | 0.2063 |  |  |
| Insurance type |  |  |  |  |
| Public | 1.50 (1.18-1.91) | 0.0011* |  |  |
| Other | 1.66 (0.91-3.02) | 0.3121 |  |  |
| ADI quintile<sup>§</sup> | 1.15 (1.04-1.27) | 0.0067* |  |  |
| Rural Address | 1.43 (1.1-1.83) | 0.0059* | 1.37 (1.05-1.78) | 0.0209* |
| Diagnoses |  |  |  |  |
| NDD | 3.57 (2.75-4.64) | <0.0001* | 2.76 (2.10-3.64) | <0.0001* |
| Seizures/Epilepsy | 1.01 (0.78-1.31) | 0.9360 |  |  |
| Headache | 0.30 (0.19-0.47) | <0.0001* |  |  |
| Number of visits<sup>§</sup> | 1.68 (1.51-1.88) | <0.0001* | 1.49 (1.32-1.68) | <0.0001* |
| Provider type |  |  |  |  |
| Specialist | 1.51 (1.11-2.06) | <0.0001* | 1.06 (0.76-1.49) | 0.2680 |
| Trainee | 2.34 (1.63-3.38) | 0.0002* | 1.90 (1.28-2.80) | <0.0001* |
| NP | 0.24 (0.11-0.54) | <0.0001* | 0.35 (0.15-0.83) | 0.0024* |
| Site of visit |  |  |  |  |
| CSCC | 0.489 (0.360-0.666) | <0.0001* |  |  |
| Other WUSTL | 0.864 (0.521-1.430) | 0.6621 |  |  |
| Combination | 1.846 (1.272-2.679) | <0.0001* |  |  |

ADI=Area Deprivation Index. CI=Confidence interval. CSCC=Children's Specialty Care Center. NDD=Neurodevelopmental disorder. NP=Nurse practitioner. OR=Odds ratio. WUSTL=Washington University in St. Louis. +Reference variables were: female sex, Black race, English language, Private insurance, Urban address, No NDD diagnosis, No seizure diagnosis, No headache diagnosis, General attending provider, and Main campus site. £ There were 10 candidate predictors for the multivariate model (age category, male sex, rural location, location of visit, insurance type, NDD diagnosis, headache diagnosis, provider type, number of visits, and ADI quintile). Five predictors were in the final multivariate model as indicated above. §Per one unit increase

### Predictors of Genetics Clinic Visit Completion in Univariate and Multivariate Analysis

To assess predictors of completion of the genetics clinic visit after referral, univariate analysis was completed for the 282 non-Hispanic Black (including multiracial) or non-Hispanic White patients who were referred to genetics clinic by neurology providers. The same independent variables were assessed as for genetics referral except language was removed due to small numbers (there were only two non-English-speaking patients among those referred to genetics, both of whom completed their genetics visits). In the univariate analysis, only race/ethnicity and number of visits were significant predictors of genetics visit completion (**Table 4**). These two variables plus provider type were included in the multivariate model based on pre-defined univariate p-value threshold of <0.1. Only race/ethnicity and number of visits remained significant in the final multivariate model. The strongest predictor of genetics visit completion was race/ethnicity, with White patients being more than twice as likely as Black patients to complete the visit (OR=2.18, 95%CI 1.06-4.48).

**Table 4:**
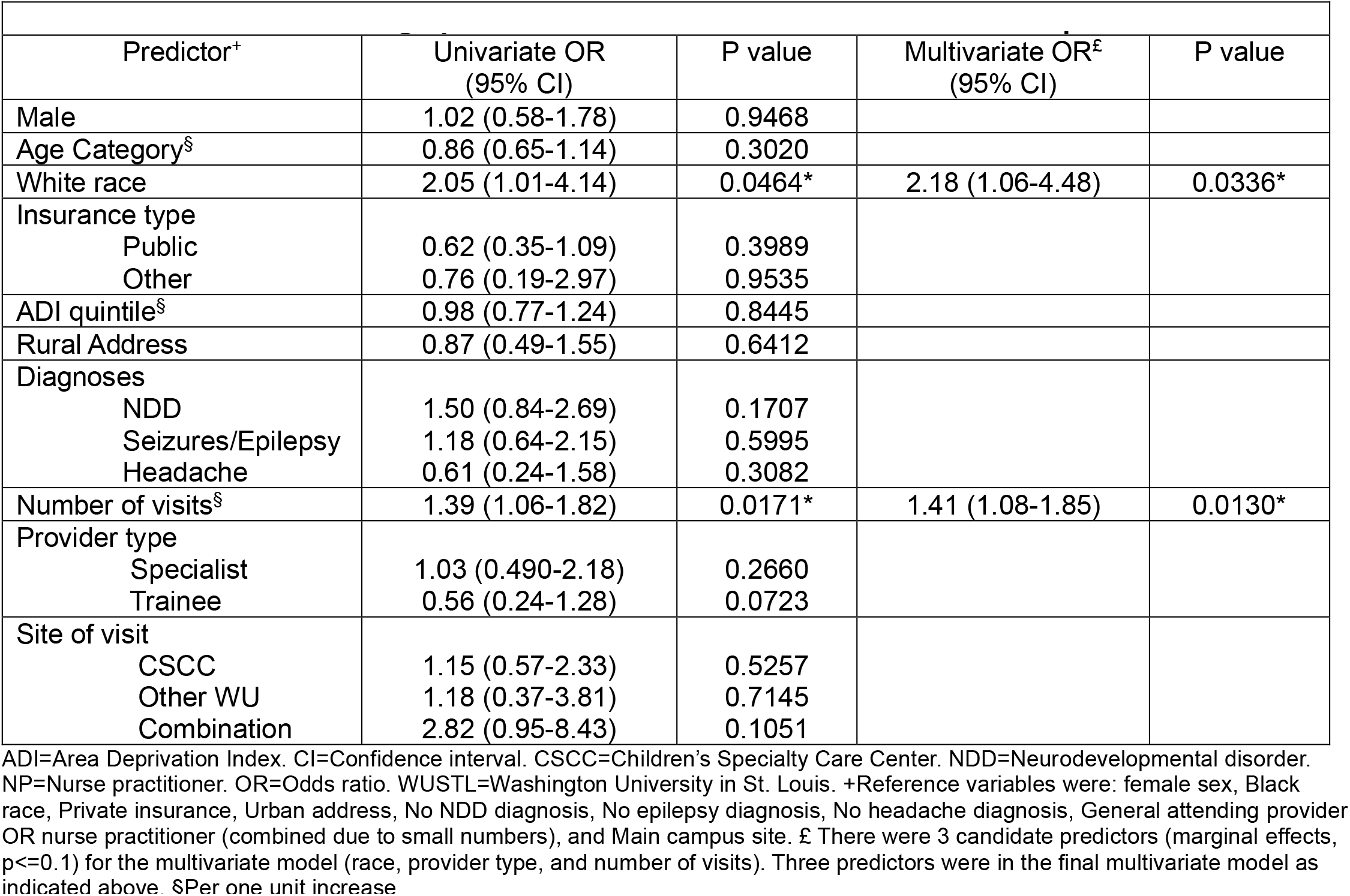
Association of Demographic & Clinical Variables with Genetic Visit Completion Status.

## Discussion

Genetic testing is an increasingly important and prevalent part of child neurology practice. We sought to explore the determinants of access to genetic testing among child neurology clinic patients at a large academic medical center, relying on an implementation science framework. In particular, we aimed to identify racial/ethnic disparities in access to genetics services, which has implications for healthcare equity, not only for diagnostics but also disease surveillance, precision therapeutics, and downstream development of novel gene therapies that are tailored toward specific gene variants that may be ancestry-specific.^39^ This paper presents the findings regarding genetics clinic referrals and completion of genetics clinic visits after referral, which represent key stages along the trajectory toward genetic testing.^16^

Several significant predictors of genetics clinic referrals and visit completion rates were identified among child neurology clinic patients. Clinical factors (patient age, NDD diagnosis, number of neurology visits) and provider-level factors (provider level of training) were more predictive of referral to genetics clinic than social factors (rural address was the only significant social determinant of referral). In contrast, regarding completion of the genetics clinic visit after referral was significantly predicted by only two factors, the number of neurology visits and race/ethnicity.

The clinical predictors of referral to genetics we uncovered were not surprising. Younger patient age was consistent with the fact that many monogenic disorders present in infancy or early childhood, and older children are more likely to present to neurology clinic for conditions such as headache/migraine, which was associated with lower likelihood of genetics referral on univariate analysis. NDD diagnosis was the strongest predictor of genetics clinic referral, which is consistent with the high rates of monogenic etiologies and academic guidelines that recommend genetic testing for all individuals with GDD/ID of unknown etiology.

Though current ACMG guidelines recommending ES/GS as first or second-line for these patients were not released until 2021 after our study dates, prior American Academy of Pediatrics guidelines recommended CMA and fragile X testing with consideration of further testing if non-diagnostic.^40^ Epilepsy/seizure diagnosis was not associated with genetics clinic referral, which may reflect neurology provider comfort with ordering and interpreting genetic testing for this indication. Number of neurology visits during the study time frame was a proxy for disease severity though it also inherently represents patient ability to access care. Attending more neurology visits within study dates was associated with a higher likelihood of referral to genetics, which is consistent with higher rates of referral for severely affected patients, in whom prior studies have demonstrated higher diagnostic yield of genetic testing compared to those who are less severely affected.^41^

Differences in genetics clinic referral based on provider type/level of training demonstrated that physician trainees were significantly more likely and NPs significantly less likely to refer compared to general attendings. These differences may relate to differences in patient cohorts, though they remained significant when controlling for NDD diagnosis, age, number of neurology visits, and rural address. Trainees may have a higher percentage of hospital follow-up or new patient visits compared to other providers, although this requires additional study. Differences in education between physicians and NPs may exist, although studies have found perceptions of inadequate genetics education in both groups.^42,43^

Surprisingly, patients from rural areas were more likely to be referred to genetics clinic than those from urban areas. Though both insurance type and ADI were significantly associated with referral in univariate analysis, with individuals on public insurance and those with higher ADI (more deprivation) more likely to be referred, but in the multivariate model these were no longer significant. Also notable is the lack of association of genetics clinic referral with patient race/ethnicity, which argues against racial/ethnic implicit bias or provider discrimination in referral pattern. This is in contrast to the adult cancer genetics literature, which found lower rates of referral among individuals identified as Black/AA or Hispanic.^44,45^ A systematic review of barriers to patient referral to genetics services, which included two pediatric studies, identified barriers including lack of patient or provider awareness of risk factors, family history, and genetic services, lack of provider knowledge about genetic conditions, inadequate coordination of referrals, and lack of genetics workforce.^46^ Unfortunately, this review did not address the impact of race/ethnicity, insurance type, or socioeconomic factors. However, they did find that awareness of genetics services was lower among health care providers practicing in rural areas and rates of referral to genetics were lower among rural providers, who were more likely to refer patients to other specialists such as oncologists compared to genetics.^46^ This may in part account for the higher rates of genetics referral among rural patients in our tertiary care suburban/urban neurology clinic population if they were less likely to have already been referred to genetics by their local (rural) health care providers.

Analysis of our other primary outcome, completion of genetics visit after referral, demonstrated only two significant predictors, one of which was also a predictor of genetics clinic referral. The shared predictor was the number of neurology visits, our marker of disease severity and access to care. Those with a greater number of neurology visits were more likely to complete their genetics visits, which is consistent with expectations that parents of more severely-affected patients may be more motivated to complete these visits, and that individuals who have demonstrated ability to access neurology care are also more able to access/complete genetics visits.

The strongest predictor of genetics visit completion was patient race/ethnicity, with White patients more than twice as likely to complete the visit compared to Black/AA patients. Reasons for this disparity remain unknown given that all other factors assessed, including patient sex, age, diagnosis, insurance type, ADI, rural/urban address, provider type, and site of visit were not associated with visit completion and thus the disparity cannot be attributed to differences in these factors between White and Black/AA patients. A study on pediatric subspecialty referrals and completion rates among a large primary and urgent care network in Pennsylvania similarly found decreased rates of visit completion among Black/AA patients, though in contrast to our findings they also saw lower rates of visit completion among those with public insurance and lower zip code median income.^19^ We suspect that structural racism plays a role in this disparity, as structural factors have led to inequities in education, economic prosperity, adverse childhood experiences, neighborhood safety, experiences of microaggressions and discrimination, and other variables not directly measured in our study.^20^ Interestingly, we did not see a racial/ethnic disparity in visit completion rate for cardiology referrals, which suggests that there may be unique differences in parent/caregiver perceptions and motivations around genetics between racial/ethnic groups. Prior research on this is very limited in pediatrics but has generally found lower genomic knowledge among Black/AA or Hispanic individuals^47,48^ but mixed results in terms of differences in perceptions of potential benefits/risks of genetic testing.^49,50^

There are several limitations to our study. First, our study was completed using EHR data, which may have inherent limitations due to the potential lack of standardization of data collection. For example, the rates of self-reported or administrative clinical staff recording of race/ethnicity are unknown. We also relied on ICD-10 and SNOMED-CT diagnostic codes, which are dependent on provider documentation and may not have complete sensitivity/specificity for the conditions studied. Another limitation was our small numbers of racial/ethnic groups other than non-Hispanic White and non-Hispanic Black. Lastly, the generalizability of our data to other clinical sites is unknown because this study data was derived from a single institution in a particular geographic region, and factors influencing genetics referrals and visit completion may vary widely based on local demographics, workforce availability, and institutional practices.

In summary, we found multiple determinants of genetics clinic referrals and completion of genetics visits among child neurology patients. We identified a racial/ethnic disparity in access to genetic services occurring at the stage of genetics visit completion after referral is placed. This suggests that incorporation of genetic testing directly into child neurology clinic and/or the establishment of multidisciplinary clinics where patients can be seen by both a neurologist and geneticist in a single visit may decrease racial disparities in access to genetic services. Further research is needed to investigate reasons behind this disparity and to develop interventions to improve access.

## Acknowledgements

The authors acknowledge the contributions of the WUSTL Center for Race, Ethnicity, and Equity and the Institute for Informatics including support from Dr. Thomas Kannampallil, PhD. The authors also acknowledge support from the WUSTL Department of Pediatrics, Division of Genetics and Genomic Medicine, including Dr. Patricia Dickson. Research reported in this publication was supported by St. Louis Children’s Hospital Foundation, the Avery Family Research Fund for Genetically Driven Developmental Disorders, National Institute of Arthritis and Musculoskeletal and Skin Diseases (R01AR06771), Washington University Institute of Clinical and Translational Sciences grant UL1 TR002345 from the National Center for Advancing Translational Sciences of the National Institutes of Health, and the Eunice Kennedy Shriver National Institute of Child Health & Human Development of the National Institutes of Health under award number P50HD103525 to the Intellectual and Developmental Disabilities Research Center at Washington University. The content is solely the responsibility of the authors and does not necessarily represent the official view of the NIH.

## Author contributions

JJC contributed to the conception and design of the study, acquisition and analysis of the data, and drafting a significant portion of the manuscript. AS contributed to the acquisition and analysis of the data. LB contributed to acquisition and analysis of the data. JH contributed to acquisition and analysis of the data. JBB contributed to the conception and design of the study. CAG contributed to the conception and design of the study. All authors contributed to editing and final approval of the manuscript.

## Data Availability

The data that support the findings of this study are available from the corresponding author, JJC, upon reasonable request.

## References

1. Manickam K, McClain MR, Demmer LA, et al. Exome and genome sequencing for pediatric patients with congenital anomalies or intellectual disability: an evidence-based clinical guideline of the American College of Medical Genetics and Genomics (ACMG). Genet Med. Nov 2021;23(11):2029–2037. doi:10.1038/s41436-021-01242-6

2. Smith L, Malinowski J, Ceulemans S, et al. Genetic testing and counseling for the unexplained epilepsies: An evidence-based practice guideline of the National Society of Genetic Counselors. J Genet Couns. Apr 2023;32(2):266–280. doi:10.1002/jgc4.1646

3. Malinowski J, Miller DT, Demmer L, et al. Systematic evidence-based review: outcomes from exome and genome sequencing for pediatric patients with congenital anomalies or intellectual disability. Genet Med. Jun 2020;22(6):986–1004. doi:10.1038/s41436-020-0771-z

4. Sheidley BR, Malinowski J, Bergner AL, et al. Genetic testing for the epilepsies: A systematic review. Epilepsia. Feb 2022;63(2):375–387. doi:10.1111/epi.17141

5. Haviland I, Daniels CI, Greene CA, et al. Genetic Diagnosis Impacts Medical Management for Pediatric Epilepsies. Pediatr Neurol. Jan 2023;138:71–80. doi:10.1016/j.pediatrneurol.2022.10.006

6. (Quality) OH. Genome-Wide Sequencing for Unexplained Developmental Disabilities or Multiple Congenital Anomalies: A Health Technology Assessment. Ont Health Technol Assess Ser. 2020;20(11):1–178.

7. McKnight D, Morales A, Hatchell KE, et al. Genetic Testing to Inform Epilepsy Treatment Management From an International Study of Clinical Practice. JAMA Neurol. Dec 01 2022;79(12):1267–1276. doi:10.1001/jamaneurol.2022.3651

8. Gonzalez-Mantilla PJ, Hu Y, Myers SM, et al. Diagnostic Yield of Exome Sequencing in Cerebral Palsy and Implications for Genetic Testing Guidelines: A Systematic Review and Meta-analysis. JAMA Pediatr. May 01 2023;177(5):472–478. doi:10.1001/jamapediatrics.2023.0008

9. Töpf A, Johnson K, Bates A, et al. Sequential targeted exome sequencing of 1001 patients affected by unexplained limb-girdle weakness. Genet Med. Sep 2020;22(9):1478–1488. doi:10.1038/s41436-020-0840-3

10. Masih S, Moirangthem A, Shambhavi A, et al. Deciphering the molecular landscape of microcephaly in 87 Indian families by exome sequencing. Eur J Med Genet. Jun 2022;65(6):104520. doi:10.1016/j.ejmg.2022.104520

11. Srivastava S, Love-Nichols JA, Dies KA, et al. Meta-analysis and multidisciplinary consensus statement: exome sequencing is a first-tier clinical diagnostic test for individuals with neurodevelopmental disorders. Genet Med. Nov 2019;21(11):2413–2421. doi:10.1038/s41436-019-0554-6

12. Srivastava S, Lewis SA, Cohen JS, et al. Molecular Diagnostic Yield of Exome Sequencing and Chromosomal Microarray in Cerebral Palsy: A Systematic Review and Meta-analysis. JAMA Neurol. Dec 01 2022;79(12):1287–1295. doi:10.1001/jamaneurol.2022.3549

13. Kutscher EJ, Joshi SM, Patel AD, Hafeez B, Grinspan ZM. Barriers to Genetic Testing for Pediatric Medicaid Beneficiaries With Epilepsy. Pediatr Neurol. Aug 2017;73:28–35. doi:10.1016/j.pediatrneurol.2017.04.014

14. Smith HS, Franciskovich R, Lewis AM, et al. Outcomes of prior authorization requests for genetic testing in outpatient pediatric genetics clinics. Genet Med. May 2021;23(5):950–955. doi:10.1038/s41436-020-01081-x

15. Penon-Portmann M, Chang J, Cheng M, Shieh JT. Genetics workforce: distribution of genetics services and challenges to health care in California. Genet Med. Jan 2020;22(1):227–231. doi:10.1038/s41436-019-0628-5

16. Lee G, Yu L, Suarez CJ, Stevenson DA, Ling A, Killer L. Factors associated with the time to complete clinical exome sequencing in a pediatric patient population. Genet Med. Oct 2022;24(10):2028–2033. doi:10.1016/j.gim.2022.06.006

17. Szego MJ, Meyn MS, Shuman C, et al. Views from the clinic: Healthcare provider perspectives on whole genome sequencing in paediatrics. Eur J Med Genet. May 2019;62(5):350–356. doi:10.1016/j.ejmg.2018.11.029

18. Kreider AR, French B, Aysola J, Saloner B, Noonan KG, Rubin DM. Quality of Health Insurance Coverage and Access to Care for Children in Low-Income Families. JAMA Pediatr. Jan 2016;170(1):43–51. doi:10.1001/jamapediatrics.2015.3028

19. Bohnhoff JC, Taormina JM, Ferrante L, Wolfson D, Ray KN. Unscheduled Referrals and Unattended Appointments After Pediatric Subspecialty Referral. Pediatrics. Dec 2019;144(6)doi:10.1542/peds.2019-0545

20. Malawa Z, Gaarde J, Spellen S. Racism as a Root Cause Approach: A New Framework. Pediatrics. Jan 2021;147(1)doi:10.1542/peds.2020-015602

21. Maina IW, Belton TD, Ginzberg S, Singh A, Johnson TJ. A decade of studying implicit racial/ethnic bias in healthcare providers using the implicit association test. Soc Sci Med. Feb 2018;199:219–229. doi:10.1016/j.socscimed.2017.05.009

22. Thompson HS, Manning M, Mitchell J, et al. Factors Associated With Racial/Ethnic Group-Based Medical Mistrust and Perspectives on COVID-19 Vaccine Trial Participation and Vaccine Uptake in the US. JAMA Netw Open. May 03 2021;4(5):e2111629. doi:10.1001/jamanetworkopen.2021.11629

23. Nolan D, Carlson M. Whole Exome Sequencing in Pediatric Neurology Patients: Clinical Implications and Estimated Cost Analysis. J Child Neurol. Jun 2016;31(7):887–94. doi:10.1177/0883073815627880

24. Augustyn M, Silver EJ, Blum N, High P, Roizen N, Stein REK. DBP Evaluations in DBPNet Sites: Is Race/Ethnicity a Significant Factor in Care? J Dev Behav Pediatr. Jan 2020;41(1):23–30. doi:10.1097/DBP.0000000000000710

25. Kiely B, Vettam S, Adesman A. Utilization of genetic testing among children with developmental disabilities in the United States. Appl Clin Genet. 2016;9:93–100. doi:10.2147/TACG.S103975

26. Woodward EN, Singh RS, Ndebele-Ngwenya P, Melgar Castillo A, Dickson KS, Kirchner JE. A more practical guide to incorporating health equity domains in implementation determinant frameworks. Implement Sci Commun. Jun 05 2021;2(1):61. doi:10.1186/s43058-021-00146-5

27. Yudell M, Roberts D, DeSalle R, Tishkoff S. SCIENCE AND SOCIETY. Taking race out of human genetics. Science. Feb 05 2016;351(6273):564–5. doi:10.1126/science.aac4951

28. Dalsgaard S, Thorsteinsson E, Trabjerg BB, et al. Incidence Rates and Cumulative Incidences of the Full Spectrum of Diagnosed Mental Disorders in Childhood and Adolescence. JAMA Psychiatry. Feb 01 2020;77(2):155–164. doi:10.1001/jamapsychiatry.2019.3523

29. Kind AJH, Buckingham WR. Making Neighborhood-Disadvantage Metrics Accessible - The Neighborhood Atlas. N Engl J Med. Jun 28 2018;378(26):2456–2458. doi:10.1056/NEJMp1802313

30. Area Deprivation Index. University of Wisconsin School of Medicine and Public Health. 2023. https://www.neighborhoodatlas.medicine.wisc.edu/

31. Rural-Urban Commuting Area Codes. Economic Research Service, U.S. Department of Agriculture. 2023. https://www.ers.usda.gov/data-products/rural-urban-commuting-area-codes/

32. RUCA Data: Using RUCA Data. Rural Health Research Center. 2023. https://depts.washington.edu/uwruca/ruca-uses.php

33. SNOMED CT to ICD-10-CM Map. March 2019 ed: National Library of Medicine; 2019.

34. Clinical Classifications Software Refined (CCSR). Accessed 2023. https://hcup-us.ahrq.gov/toolssoftware/ccsr/ccs_refined.jsp#overview

35. SAS System for Windows. Version 9.4. SAS Institute Inc; 2013.

36. Fox J G. M. Generalized collinearity diagnostics. J AmStat Assoc. 1992;87(417):178–183.

37. Profile of General Population and Housing Characteristics: St. Louis, MO-IL Metro Area. 2020. https://data.census.gov/table?g=310XX00US41180&d=DEC+Demographic+Profile&tid=DECENNIALDP2020.DP1

38. Hispanic or Latino, and Not Hispanic or Latino by Race. 2020. https://data.census.gov/table?q=race+and+ethnicity&g=040XX00US29&y=2020&tid=DECENNIALPL2020.P2

39. McGarry ME, McColley SA. Cystic fibrosis patients of minority race and ethnicity less likely eligible for CFTR modulators based on CFTR genotype. Pediatr Pulmonol. Jun 2021;56(6):1496–1503. doi:10.1002/ppul.25285

40. Moeschler JB, Shevell M, Genetics Co. Comprehensive evaluation of the child with intellectual disability or global developmental delays. Pediatrics. Sep 2014;134(3):e903–18. doi:10.1542/peds.2014-1839

41. Evers C, Staufner C, Granzow M, et al. Impact of clinical exomes in neurodevelopmental and neurometabolic disorders. Mol Genet Metab. Aug 2017;121(4):297–307. doi:10.1016/j.ymgme.2017.06.014

42. Maradiegue AH, Edwards QT, Seibert D. 5-years later - have faculty integrated medical genetics into nurse practitioner curriculum? Int J Nurs Educ Scholarsh. Oct 31 2013;10doi:10.1515/ijnes-2012-0007

43. Sen K, DiSabella MT, Harmon J, Strelzik JA, Gropman AL. Gaps in Neurogenetics Education During Child Neurology Residency: Results of a National Survey. J Child Neurol. Aug 2022;37(8-9):702–706. doi:10.1177/08830738221106896

44. Muller C, Lee SM, Barge W, et al. Low Referral Rate for Genetic Testing in Racially and Ethnically Diverse Patients Despite Universal Colorectal Cancer Screening. Clin Gastroenterol Hepatol. Dec 2018;16(12):1911–1918.e2. doi:10.1016/j.cgh.2018.08.038

45. Ademuyiwa FO, Salyer P, Tao Y, et al. Genetic Counseling and Testing in African American Patients With Breast Cancer: A Nationwide Survey of US Breast Oncologists. J Clin Oncol. Dec 20 2021;39(36):4020–4028. doi:10.1200/JCO.21.01426

46. Delikurt T, Williamson GR, Anastasiadou V, Skirton H. A systematic review of factors that act as barriers to patient referral to genetic services. Eur J Hum Genet. Jun 2015;23(6):739–45. doi:10.1038/ejhg.2014.180

47. Rini C, Henderson GE, Evans JP, et al. Genomic knowledge in the context of diagnostic exome sequencing: changes over time, persistent subgroup differences, and associations with psychological sequencing outcomes. Genet Med. Jan 2020;22(1):60–68. doi:10.1038/s41436-019-0600-4

48. Lewis KL, Heidlebaugh AR, Epps S, et al. Knowledge, motivations, expectations, and traits of an African, African-American, and Afro-Caribbean sequencing cohort and comparisons to the original ClinSeq. Genet Med. Jun 2019;21(6):1355–1362. doi:10.1038/s41436-018-0341-9

49. Palmer CG, Martinez A, Fox M, Sininger Y, Grody WW, Schimmenti LA. Ethnic differences in parental perceptions of genetic testing for deaf infants. J Genet Couns. Feb 2008;17(1):129–38. doi:10.1007/s10897-007-9134-z

50. Tarini BA, Singer D, Clark SJ, Davis MM. Parents’ interest in predictive genetic testing for their children when a disease has no treatment. Pediatrics. Sep 2009;124(3):e432–8. doi:10.1542/peds.2008-2389

